# Assessing the impact of coordinated COVID-19 exit strategies across Europe

**DOI:** 10.1101/2020.06.16.20132688

**Authors:** N.W. Ruktanonchai, J.R. Floyd, S. Lai, C.W. Ruktanonchai, A. Sadilek, P. Rente-Lourenco, X. Ben, A. Carioli, J. Gwinn, J.E. Steele, O. Prosper, A. Schneider, A. Oplinger, P. Eastham, A.J. Tatem

## Abstract

As rates of new COVID-19 cases decline across Europe due to non-pharmaceutical interventions such as social distancing policies and lockdown measures, countries require guidance on how to ease restrictions while minimizing the risk of resurgent outbreaks. Here, we use mobility and case data to quantify how coordinated exit strategies could delay continental resurgence and limit community transmission of COVID-19. We find that a resurgent continental epidemic could occur as many as 5 weeks earlier when well-connected countries with stringent existing interventions end their interventions prematurely. Further, we found that appropriate coordination can greatly improve the likelihood of eliminating community transmission throughout Europe. In particular, synchronizing intermittent lockdowns across Europe meant half as many lockdown periods were required to end community transmission continent-wide.

**One Sentence Summary:** EU coordination in easing restrictions is key to preventing resurgent COVID-19 outbreaks and stopping community transmission.

---

The ongoing COVID-19 pandemic rapidly spread across Europe throughout February and March 2020, making it the largest cluster of cases worldwide (*1*). In response, most of Europe implemented strict lockdown measures to control disease spread, which have been shown to be effective at reducing transmission (*2*–*4*). As rates of new cases decline, countries are now implementing various exit strategies to relax restrictions (*5*). Long-term success of any potential exit strategy hinges on what happens regionally, as international importation could overwhelm efforts to prevent resurgence through testing and contact tracing (*6, 7*). To account for this, the European Commission recommended that governments provide advance warning of plans to relax non-pharmaceutical interventions (NPIs) (*8*), and in particular, has focused on coordinated easing of travel restrictions (*9*). To better inform the importance and nature of an internationally coordinated exit strategy, governments require an evidence base for understanding importation and the consequences of easing interventions in an uncoordinated way.

Data from mobile phones can help address this by informing connectivity patterns, contact rates, and the effect of various NPIs on mobility. In other settings, they have been instrumental for understanding where infection occurs for various diseases (*10*) such as malaria (*11, 12*), predicting disease spread (*13*), and quantifying population mobility during and after catastrophic events (*14*). More recently, for the COVID-19 pandemic, mobile phone data have been valuable in assessing NPI effectiveness (*4, 15*), and remain at the forefront of understanding whether populations are adhering to social distancing policies (*16*– *19*). These data link well with theoretical models that provide a basis for understanding how heterogeneous mobility and exposure will affect disease invasion (*20*) in spatially structured populations (*21*), as well.

Here we provide an evidence base for coordinated exit strategies across Europe using mobile phone data and a metapopulation model of COVID-19 transmission (*22*). Specifically, we quantify the progression of a second epidemic continent-wide if countries act in a coordinated or uncoordinated manner. We also quantify how coordination could influence regionally interrupted transmission of COVID-19, testing the importance of synchronized NPIs if countries phase them to limit economic impact. We accomplished this by 1) estimating pre-COVID-19 mobility using a novel anonymized and aggregated call data record (CDR) dataset from Vodafone and an anonymized and aggregated continental NUTS3 (Nomenclature of Territorial Units for Statistics) mobility dataset from Google (Table S1), 2) measuring mobility reductions due to NPIs using a separate COVID-19 Google dataset, and 3) propagating these reductions in an epidemiological model (see Fig S1 for data flow). All analyses were undertaken at the NUTS3 administrative unit level, which are administrative boundaries regulated by the European Union (EU) for use within EU member states (35), with spatial extents defined by population thresholds ranging between 150,000 to 800,000 residents.

First, we predicted the baseline probability of moving between NUTS3 regions across Europe using the Vodafone data in Spain and Italy and the continental Google NUTS3 dataset (Fig 1).

**Fig. 1.**
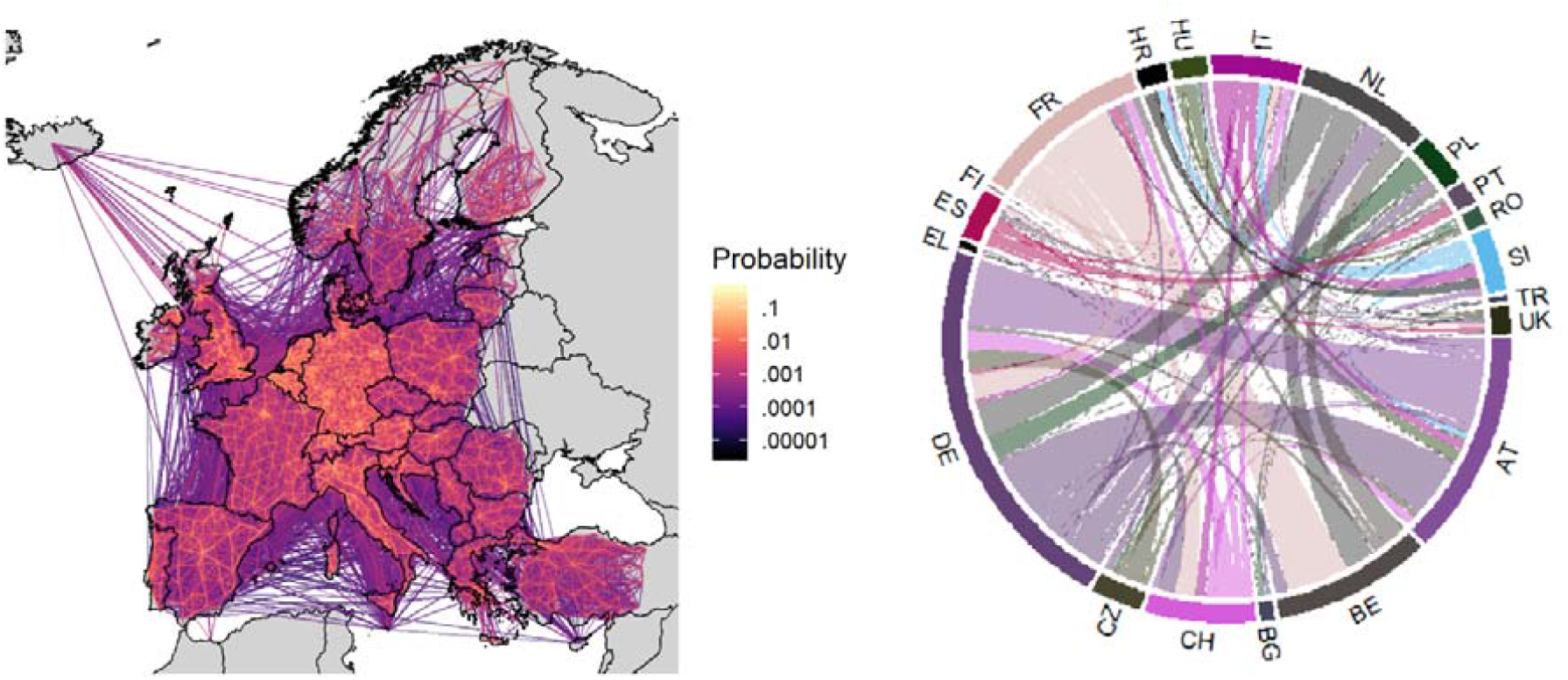
Predicted baseline mobility patterns for Jan 28 – Feb 18, 2020. (Left) Probability of people moving between NUTS3 administrative units per 8 hours. (Right) Individual probability of moving between top 20 European countries with the greatest outward mobility. For example, an individual in Germany (DE) is roughly twice as likely to travel internationally as compared to an individual in Austria (AT). Colors shown in the left panel correspond to the source country, and country codes shown are from Eurostat (*46*).

We then analyzed the Google COVID-19 dataset to quantify reductions in mobility and contact rates from January 2020 through the end of March 2020 in response to the COVID-19 pandemic (Fig 2). In our simulations, we used observed reductions in mobility in each NUTS3 area to proportionally reduce outgoing flows, incoming flows, and local contact rates for that area.

**Fig. 2.**
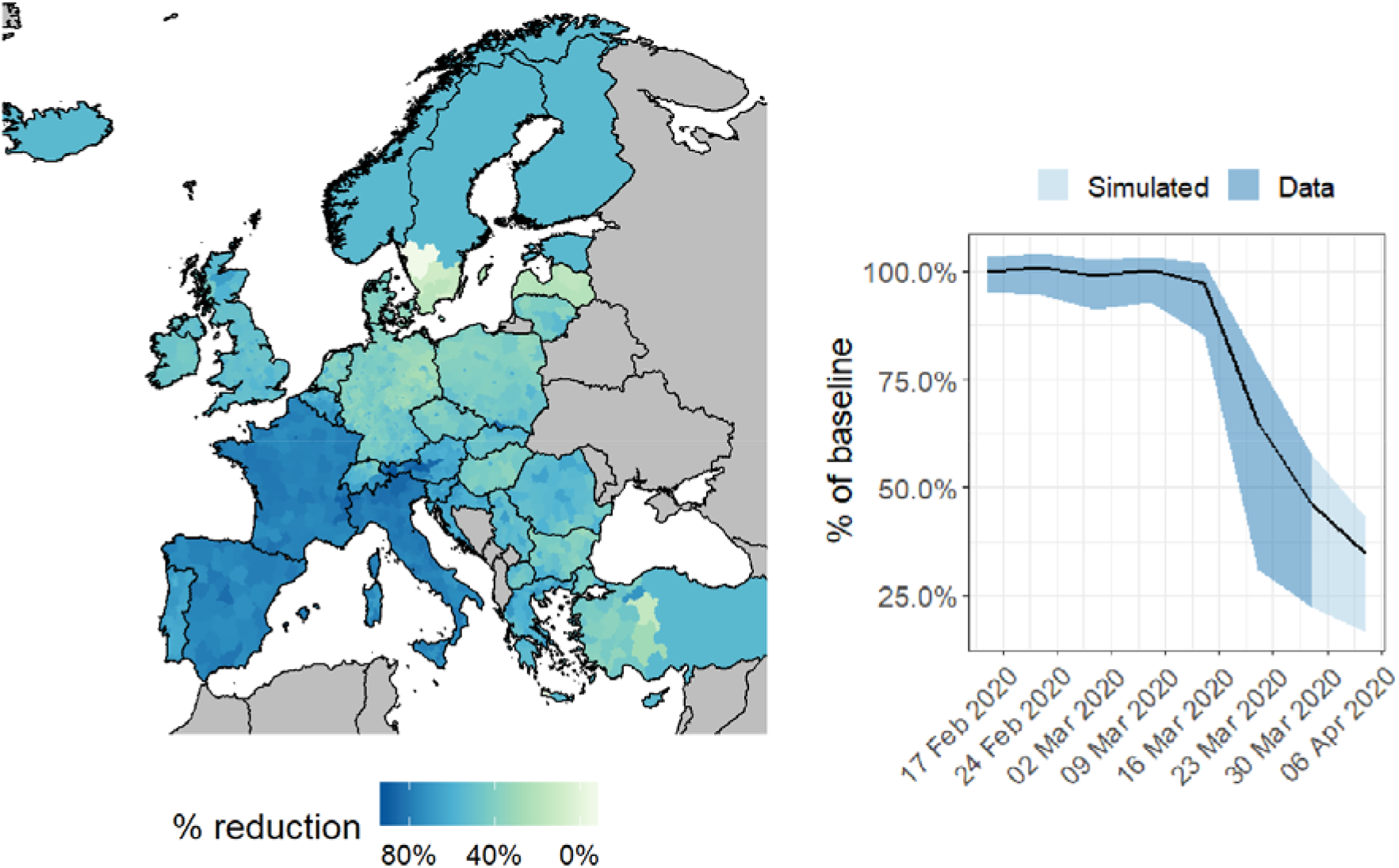
Reduction in mobility observed in NUTS3 areas from Feb 11 to Apr 6, 2020. (Left) Reduction in mobility observed in each NUTS3 administrative unit across Europe for the week of Mar 21-28, 2020 compared to Jan-Feb 2020. Movement data were not available for countries in grey. (Right) Weekly average change in mobility across all NUTS3 areas. Dark blue shows reductions observed in the Google COVID-19 dataset, light blue shows extrapolation of reductions by one week. Black line shows mean change compared to baseline. When implementing NPIs in various NUTS3 areas, we used the movement reduction estimates for the end of this period, Apr 6 2020.

Using these baseline mobility patterns and reductions in mobility, we simulated the spread of COVID-19 over 6 months starting April 4, 2020 while making various assumptions about where and when NPIs would be relaxed or reinstated. Across all simulations, we started transmission on March 20, 2020 because this predates large reductions in mobility (Fig 2; right), allowing the disease to spread initially in a data-driven way that can help account for spatial biases in reporting and testing. We parameterized initial numbers of people infected using a repository maintained by the Johns Hopkins University CSSE (*23*). Because the case data from this repository were country-level, we distributed cases across NUTS3 area proportionally based on population size (Fig S10).

To simulate different exit strategies and the overall impact of different NPIs enacted, we reduced mobility one week past March 28 (for March 29 – April 4) based on the observed change between March 15 – 21 to March 22 – 28 to account for changes caused by further uptake of existing NPIs (Fig 2). On average, this represented an overall mean 65% reduction in mobility compared to January 28 – February 18, agreeing with recent studies on contact rate reductions in the UK (*24*), which observed a 73% reduction in daily contacts. When simulating active lockdowns on dates past April 4, we used the predicted mobility reduction for each NUTS3 area from March 29 – April 4. When we simulated countries lifting their NPIs entirely, we used the relative mobility patterns observed during March 1 – 7.

First, we compared secondary epidemic timing when all countries coordinated their exit strategies with simulations where one country ended their interventions early. We iteratively tested the impact of each country in Europe easing lockdowns starting April 15, while all other countries extended their NPIs for 4, 8, and 12 weeks, depending on simulation run. For the country that lifted their NPIs early, we assumed people in each NUTS3 area would voluntarily reduce their average contact rate by 20% compared to the January – February baseline, or slightly less than the reduction in mobility observed on March 23, because countries that have lifted NPIs have observed sustained limited mobility reductions beyond the relaxing of various restrictions (17).

If a country lifted their NPIs early, we found a second epidemic could occur much earlier (Fig 3; left). The right panel of Figure 3 measures the earlier timing to 25% of people across Europe having had COVID-19 (infected + recovered + exposed; see Fig S13 for plot showing this explicitly). This measure captures when uncontrolled widespread transmission occurred while accounting for multiple peaks and varying peak heights in Fig 3L. Time to 25% infected was particularly sensitive to well-connected countries that implemented strong NPIs, such as France and Italy (Fig 3; right). France lifting their NPIs early led to the earliest second epidemic, 35 days earlier than if all countries lifted their NPIs simultaneously (interquartile range from 32.3 to 36.8 days). Despite having experienced relatively low reductions in mobility through March 28, Germany remains important to continental resurgence, due to high connectivity with neighboring countries (Fig 1; right). When exploring the epidemic curves through time when different countries lifted their NPIs early, we found that different types of mobility initiated continental epidemics. While France lifting their NPIs early led to resurgence in major population centers continent-wide, Germany lifting NPIs early led to resurgence in neighboring countries first (Fig S15). Further, certain areas keeping R slightly over 1 under NPIs also led to an initial peak in some simulations, and maintained the threat of resurgence even after 12 weeks of NPIs continent-wide (Fig 3, left; see SI section “Exploring spatiotemporal dynamics of spread” for more detail).

**Fig. 3.**
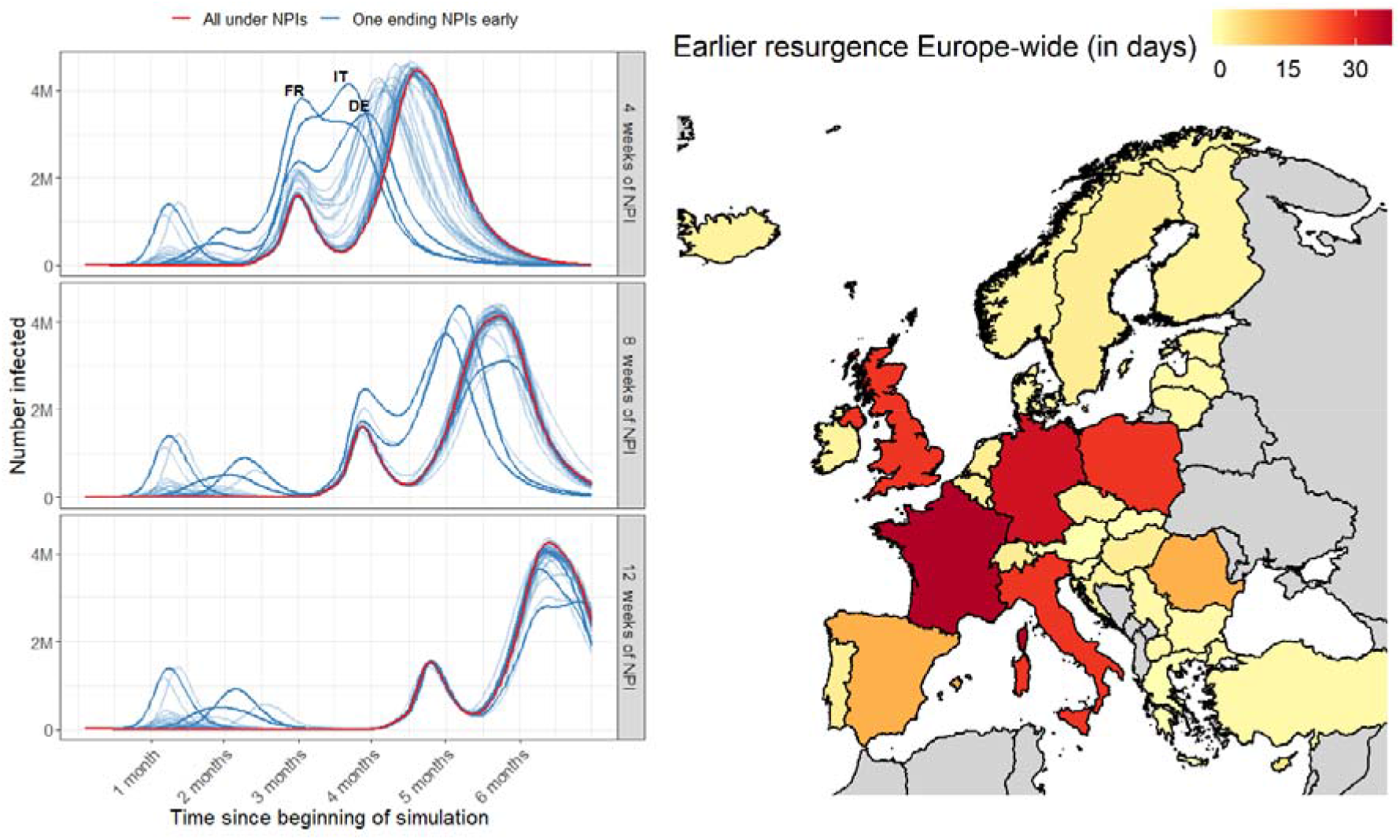
Epidemic spread if all countries but one maintain existing NPIs. When lifting NPIs early, countries reverted to baseline mobility on April 15. (Left) Epidemic curves, with varying numbers of weeks that NPIs are implemented. Curves indicate numbers of active cases at any given time, rather than numbers of new cases per day. Red lines indicate epidemic curves where all countries maintain NPIs for the denoted number of weeks. Blue lines indicate epidemic curves if one country ends intervention policies early (each line represents one randomly chosen country that ends its policies early); France, Germany, and Italy are highlighted. (Right) For the 4 weeks of NPI scenario, the number of days earlier that an uncontrolled second epidemic occurs continent-wide if each country ends NPIs early, measured as the time to 25% of the population of Europe having had COVID-19. Movement data were not available for countries in grey.

We also tested how cycling NPIs in a synchronized or unsynchronized way affected the continent-wide epidemic. Cycled NPIs meant countries switched between being under interventions for several weeks and under no interventions for the same number of weeks over a number of cycles. Synchronized NPIs meant all countries implemented lockdowns at the same time, while unsynchronized NPIs meant half of all countries (randomly chosen for each simulation run) were under lockdown at any time. Cycling NPIs reflects the intermittent lockdowns that could occur if countries reinstate interventions after surpassing threshold numbers of new cases (*25, 26*). Therefore, this test helps predict what may happen if countries do not coordinate the easing and reinstating of NPIs based on regional rates of new cases.

Across 1200 simulations, we found that synchronized cycles of NPIs were always more likely to end community transmission over 6 months, and generally lowered transmission further than if NPIs were unsynchronized (Fig 4). In the most striking example, four synchronized cycles were enough to eliminate local COVID-19 cases in 90% of simulations, while four unsynchronized cycles only led to elimination 5% of the time. Two synchronized cycles of four-week NPIs were also sufficient to end community transmission, whereas four unsynchronized cycles of four-week NPIs were necessary to end community transmission (Fig 4, right). The only simulations where unsynchronized NPIs had fewer cases than synchronized NPIs at the end of simulation was with 2 cycles of 3 week-long NPIs (Fig 4, top left), which occurred because enough people were infected under unsynchronized NPIs that herd immunity reduced transmission.

**Fig. 4.**
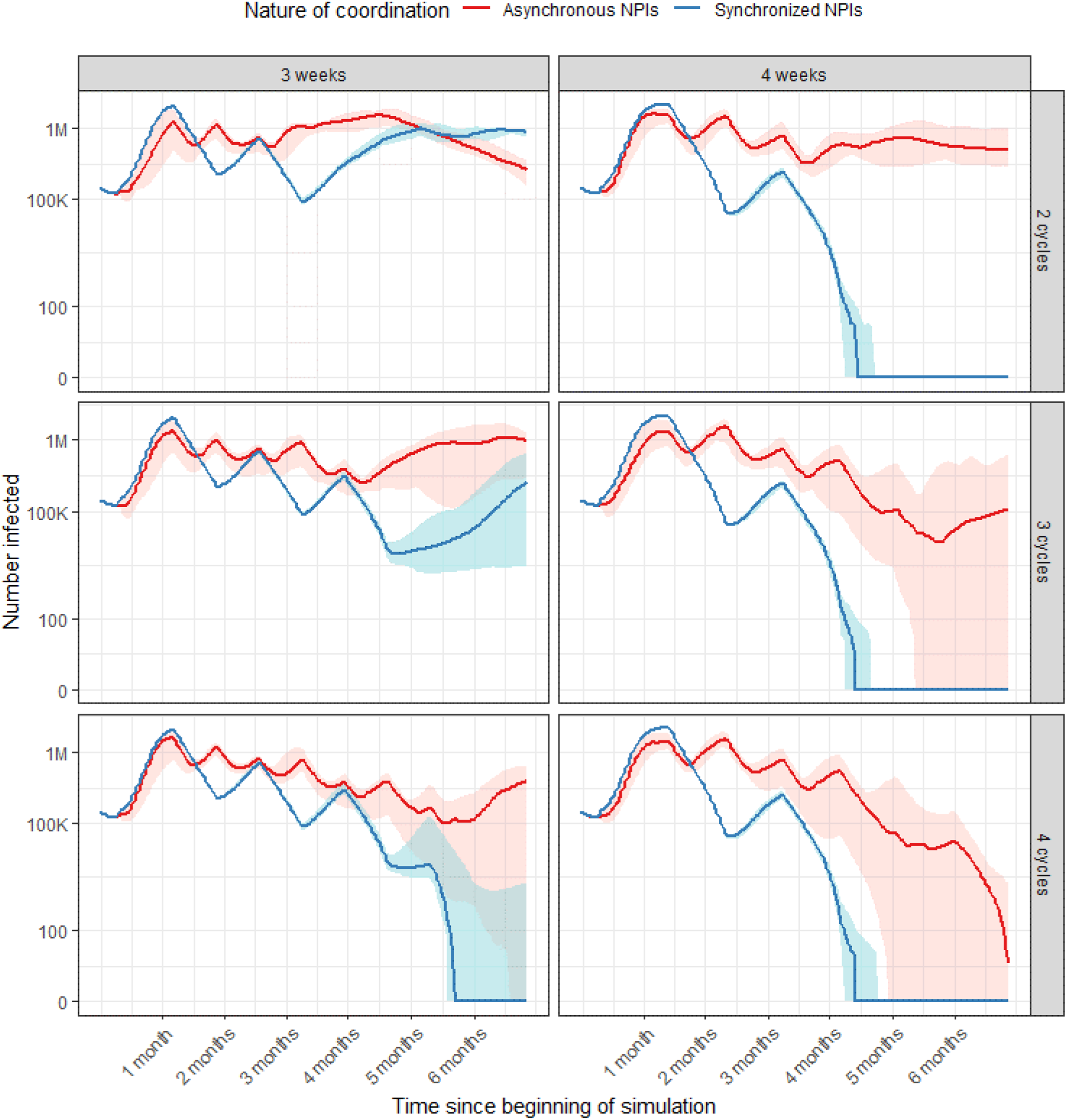
Cases over time, when NPIs are synchronized and unsynchronized across all European countries. Rows vary the number of on-off cycles that occur, and columns indicate the number of on-off cycles implemented. Red: Cases when European countries do not synchronize NPI timing. Blue: Cases when European countries are all synchronized in NPI timing. Shaded areas indicate intervals in which 95% of simulations fell within, over 200 simulations.

Intergovernmental organizations such as the World Health Organization have stressed the importance of international solidarity in terms of sharing resources and expertise in combating COVID-19 (*1*). Our results reiterate this, as one country ending NPIs before others could mean disease resurgence across Europe as many as 5 weeks earlier, reducing the time available to expand test-and-treat and to develop new therapeutics or vaccines (Fig 3).

Heterogeneities in mobility reduction (Fig 2), baseline mobility patterns (Fig 1), and population sizes mean that certain countries are particularly important to continental resurgence, such as France, Germany, Italy, and Poland (Fig 3).

These key countries varied in how local cases led to continental resurgence, implying different key interventions for each. For example, while spread out of Germany led to epidemics in neighboring countries initially, spread out of France led to epidemics in population centers continent-wide (Fig S12). This suggests that continued airport closures could be key for preventing spread from France to other countries across Europe, while in Germany, limiting local travel may be more effective.

Further, we found that small pockets of community transmission under NPIs could assure a second epidemic wave continent-wide. In our study, this occurred due to central Turkey experiencing limited mobility reductions (Fig 2) and exhibiting a high starting reproductive number (Fig S2) that kept local R slightly above 1 when under NPIs. This had dramatic effect across many simulations, as in simulations where small countries lifted their NPIs early, we found that resurgence was largely driven by importation from central Turkey and exhibited epidemic curves very similar to the case where all countries maintained NPIs (Fig 3). While the actual mobility reduction in central Turkey is uncertain and has likely changed since late March 2020, this highlights the importance of countries ensuring that R stays below 1 during lockdown periods, and the importance of effective screening of international travelers from areas with sustained transmission well into the future.

We also found that the nature of coordination was key to reducing resurgence risk. When cycling NPIs between weeks, synchronized interventions across all countries meant that cases could be driven down more quickly (Fig 4). Fewer cases at the end of the synchronized lockdowns led to much higher likelihoods of reaching zero cases locally, due to a higher chance of stochastic recovery processes leading to interrupted transmission. In real terms, the synchronized scenario approximates what could happen if countries set case thresholds for lifting NPIs regionally, while the unsynchronized scenario simulates what could happen if countries only consider case numbers within their boundaries.

This study has several limitations that influence the direct applicability of our case number predictions across Europe. First, we use observed mobility reductions as a proxy for reductions in contact rate, which may not reflect reality, though the contact rate reduction estimates from this process accord with those observed in other studies (*24*). Secondly, COVID-19 is known to exhibit age-dependent severity, and contact rates are strongly age-dependent (*27, 28*), which could introduce heterogeneities that we did not incorporate into our simulations. We believe our results should be robust to these limitations, however, because we ran our simulations over different values of *R* and varying the serial interval. In these sensitivity analyses, we found that the existence of key countries and the importance of coordinated NPIs were robust to these changes (see SI), though varying the serial interval could extend or shrink the second epidemic timings and epidemic peaks observed in Fig 3. Additionally, we reduced mobility for each NUTS3 area uniformly. In reality, long-distance movement reduced much more than short-distance movement due to country-level travel restrictions and other NPIs (Fig S8). This likely provides a continental protective effect against early resurgence compared to our results, particularly if quarantining measures are put in place for long-distance travelers.

Our mobility estimates may also be biased due to the populations included in the Google and Vodafone data. Google’s consumer Location History feature is only available from smartphone users, is turned off by default, and is viewed through the lens of differential privacy algorithms designed to protect user privacy and obscure fine detail. Vodafone’s anonymized and aggregated data were based on network data from customers who had full control over their privacy settings, potentially introducing biases as well. This work makes a step towards using multiple datasets to capture population-level patterns that go beyond any one service or system. Further, because both the Google and Vodafone data are aggregate datasets, we could not account for individual-level correlation in mobility patterns in our model (i.e. individuals who travel elsewhere but return home shortly thereafter). This likely means our model will overestimate spread and resurgence in general, as infectious people will end up less likely to return home.

Coordination will be key to an effective, equitable response to COVID-19. This means not just sharing resources, but also ensuring that exit strategies account for neighboring countries and regions. While coordinating exit strategies across an entire continent may prove politically difficult, the presence of key countries and community structure offer possible coordination groups that do not require engagement from all countries (see SI, community detection). Further, coordinated exit strategies that account for real-time case data will likely improve outcomes compared to our predictions, as we simulated intermittent NPIs that were lifted regardless of actual transmission context. A multifaceted, reactive approach to lifting NPIs will be necessary to minimize resurgence risk. This means beyond international cooperation, robust test-and-treat (*30*) and household quarantine (*31*) measures should be in place. Future work will further inform the role mobility, NPIs, and international coordination can play in slowing COVID-19 resurgence, building on existing work (*29*) examining invasion, re-invasion, and disease extinction (*32*) in spatially structured populations.

The implications of our study extend well beyond Europe and COVID-19, broadly demonstrating the importance of communities coordinating easing of various NPIs for any potential pandemic. In the United States, NPIs have been generally implemented at the state-level, and because states will be strongly interconnected, our results emphasize national coordination of pandemic prepararedness efforts moving forward. Elsewhere, relatively porous national borders between many lower and middle income countries mean without coordination, these countries may have to deal with significant international importation after controlling local transmission (*33*). COVID-19 transmission and transmission of any infectious disease will ignore national and provincial borders; preventing resurgence and spread will mean ensuring that pockets of transmission do not persist in areas with limited interventions at the expense of later epidemics in others.

## Data Availability

ERGO ID 48113

## Acknowledgments

The authors would like to acknowledge JP, SW, EF and the DG group for insightful discussions on heterogeneously applied interventions.

## Author contributions

NWR, JRF, SL, PRL, AJT designed the research. NWR, JRF, SL, CWR, PRL, OP, AC built the model and ran simulations. NWR, JRF, SL, CWR, AS, PRL, AC, carried out analyses. AS, PRL, XB, AC, JES, AS, AO, PE provided technical support. AS, PRL, XB, AS, AO, PE helped with data curation. AS and XB created the Google COVID-19 aggregated dataset. NWR, JRF, SL, CWR, AC, AS, PRL, JG, OP, AJT wrote and edited the manuscript.

## Data and materials availability

Code for the model simulations is available at the following GitHub repository: https://github.com/wpgp/BEARmod. doi:10.5281/zenodo.3875099. The data on COVID-19 cases and interventions reported by country are available from the data sources listed in Supplementary Materials. The original population movement data obtained from Google and Vodafone for this study are not publicly available since this would compromise the agreement with the data provider, but data requests should be made to the corresponding authors.

